# Current quantitative polymerase chain reaction to detect severe acute respiratory syndrome coronavirus 2 may give positive results for other described coronavirus

**DOI:** 10.1101/2021.02.08.21251332

**Authors:** Antonio Martínez-Murcia, Adrián García-Sirera, Aaron Navarro, Patricia Ros-Tárraga, Laura Pérez

## Abstract

Some weeks after the first CoVID-19 outbreak, the WHO published some qPCR protocol assays developed by different institutions worldwide. These qPCR designs are being used to detect the presence of SARS-CoV-2 in the population, which allow us to monitore the prevalence of the virus during the pandemic. Moreover, the use of these designs is wide spreading and nowadays they are used to detect SARS-CoV-2 in environmental samples to act as epidemiological surveillance tool. However, at the time of designing the published RT-qPCR assays, a lack of SARS-CoV-2 genomes available may explain a low exclusivity in some cases. In this study, we are reporting experimental data which demonstrate that some of the current qPCR used to detect SARS-CoV-2 may give positive results for other described coronavirus different from SARS-CoV-2.

## INTRODUCTION

The Coronavirus Disease 2019 (CoVID-19), characterized as a pandemic by the World Health Organization (WHO), is caused by the Severe Acute Respiratory Syndrome Coronavirus 2 (SARS-CoV-2), a *Betacoronavirus* with similar characteristics than SARS-CoV, Bat SARS-CoV and Pangolin CoV, among others (Ceraolo et al, 2020; Zhou et al., 2020). Today, 8^th^ February 2021, the number of CoVID-19 positive cases globally surpass 105.3 million people, with more than 2.3 million deaths (WHO, 2020a).

At the beginning of the outbreak, WHO supported access to SARS-CoV-2 in-house polymerase chain reaction (PCR) protocol assays by posting them online on the WHO website. These real-time PCR or quantitative PCR (qPCR) designs were developed by different institutes worldwide: Charité Virology, Berlin, Germany (E and RdRp with P1 and P2 probes, last one exclusive for SARS-CoV-2); Institut Pasteur, Paris (IP2 and IP4); Centers for Disease Control and Prevention (CDC), Division of Viral Diseases, Atlanta, USA (N1 and N2); National Institute for Viral Disease Control and Prevention (CDC), China (ORF1ab and N); Hong Kong University (ORF1b and N); Department of Medical Sciences, Ministry of Public Health, Thailand (N); the National Institute of Infectious Diseases, Japan (N) (WHO, 2020b). Due to the availability of the oligo sequences and the support from prestigious organizations, these qPCR designs were the basis to develop qPCR methods and many commercial qPCR kits. Most of them are being used by laboratories worldwide to detect the presence of SARS-CoV-2 in human samples, but also from the environment, like wastewater samples. RT-qPCR tests in population are a short-term solution to assess the prevalence of the virus in a determinate date, but setting up feasible and reliable methods for SARS-CoV-2 epidemiological surveillance is necessary to successfully combat this pandemic virus and apply efficient preventive measures in case of virus reappearance. Difficulties of an easy symptom-based diagnosis (particularly at early stages of the infection), the existence of a considerable asymptomatic population, the lack of effective treatment or vaccine, and the widespread nature of the pandemics, have enforced radical and extremely costly epidemiological control measures, including worldwide lockdowns.

Scientific reports have demonstrated that asymptomatic and recently recovered patients can excrete SARS-CoV-2 in faeces and urine (Cheung et al., 2020; Lescure et al., 2020; Lo et al., 2020; Randazzo et al., 2020a; Sun et al., 2020; Wang et al., 2020). Also, the fact that SARS-CoV-2 can be transmitted by two ways, directly through respiratory droplets and indirectly through fomites, is accepted because positive results were obtained from samples of various environmental surfaces, air and sewage in hospital and community settings (Mouchtouri et al., 2020; WHO, 2020c). All these together have generally been taken as support to consider that environmental samples from different places, like those from wastewater, could act as an effective epidemiological surveillance tool. However, when testing SARS-CoV-2 by qPCR in environmental samples, the exclusivity of primers/probe sequences becomes extremely relevant because the diversity of organisms in nature is huge when compared with human clinical samples, where only some selected SARS-CoV-2 clones are expected. In a previous paper, we analysed the *in silico* exclusivity of some of the most worldwide used qPCR designs published by WHO and our own qPCR design transferred as *GPS™ CoVID-19 dtec-RT-qPCR Test* and commercially available (MartÍnez-Murcia et al., 2020). The analysis showed a relatively low SARS-CoV-2 *in silico* specificity in several of the qPCR designs, while the qPCR available by GPS™ was the most exclusive by far. However, experimental work supporting above analysis was not available at that time.

In the present study, we empirically demonstrated that some of the currently qPCR used to detect SARS-CoV-2 may give positive results for other described coronavirus. This finding is supported by experimental results of qPCR assays using synthetic sequence templates belonging to a non-SARS-CoV-2 coronavirus.

## MATERIALS AND METHODS

The primers and probes sequences of the qPCR designs subjected to analysis in the present study (CDC Atlanta, 2020; Corman et al., 2020; Institut Pasteur, 2020; MartÍnez-Murcia et al., 2020) were used to find similar regions between close biological sequences using the Basic Local Alignment Search Tool (BLAST) software available on the National Center for Biotechnology Information (NCBI, https://blast.ncbi.nlm.nih.gov/Blast.cgi) website databases (Bethesda, MD, USA). The closest coronavirus strain for each one, different from SARS-CoV-2, was selected and used to test their *in vitro* exclusivity.

In order to test the *in vitro* exclusivity of the qPCR designs, different synthetic DNA templates provided by IDT (California, USA) and Eurofins Genomics Germany GmbH (Ebersberg, Germany) were used. They were synthetised containing full match sequences, to act as a positive control for each qPCR design, and the corresponding target sequence of the selected Pangolin CoV strain. A non-related target control sequence was added to synthetic templates to calibrate template copy number. Primers and probes of IP2 and IP4 designs were provided by IDT (California, USA), and these of RdRp-P2, N1 and N2 were obtained from Eurofins Genomics Germany GmbH (Ebersberg, Germany). Finally, *GPS™ CoVID-19 dtec-RT-qPCR Test* was supplied by Genetic Analysis Strategies SL (Alicante, Spain).

Synthetic templates were diluted by preparing 10-fold dilution series containing 10 to 10^6^ copies of synthetic templates. qPCR tests, were performed following the recommended protocols by the corresponding research institute, using the GPS™ LyoMix RT-qPCR (Alicante, Spain). As recommended for each qPCR design, we used an amplification step of 50 cycles at 95°C for 15 s and at 58°C for 30 s for IP2 and IP4 (Institut Pasteur, 2020); 45 cycles at 94°C for 15 s and at 58°C for 30 s for RdRp-P2 (Corman et al., 2020); and 45 cycles at 95°C for 3s and at 55°C for 30 s for N1 and N2 (CDC Atlanta, 2020). Finally, the GPS™ *CoVID-19 dtec-RT-qPCR Test* was subjected to qPCR following the manufacturer’s instructions (MartÍnez-Murcia et al., 2020). Reverse transcription and activation steps were performed, according to the LyoMix RT-qPCR used, at 50°C for 10 min and 95°C for 1 min. Positive and negative PCR controls were included, and reaction mixtures were subjected to qPCR in a StepOne thermocycler (Applied Biosystems, USA).

## RESULTS AND DISCUSSION

The alignments performed between primers and probes sequences of the qPCR designs published by the WHO and the GPS™ qPCR kit, showed that Pangolin CoV isolate MP789, complete genome (Genbank No.: MT121216.1) was the closest virus sequence for IP4 and N1 qPCR designs; and Bat coronavirus RaTG13, complete genome (Genbank No.: MN996532.1) for IP2, N2, E, RdRp-P2 and GPS™ kit. RaTG13 is a single Bat-CoV sequence which shows the highest homology (96.7%) with all the SARS-CoV-2 sequences, and it was deposited the 27^th^ January, much after the outbreak started. Therefore, the possibility of RNA contamination during genome sequencing should be ruled-out before taking further conclusions (MartÍnez-Murcia et al., 2020). For this reason, the closest viral sequences selected were Pangolin CoV isolate MP789, complete genome (Genbank No.: MT121216.1) for IP2, E and RdRp-P2 qPCR designs; Pangolin CoV isolate PCoV_GX-P2V, complete genome (Genbank No.: MT072864.1) for N2; and hCoV-19/pangolin/Guangxi/P1E/2017 (Accesion ID: EPI_ISL_410539) for GPS™ qPCR kit. Information about the target sequence on Pangolin CoV sequence selected of each qPCR design and the amplicons size are summarized in Table 1.

**Table 1.** Summary of the coronavirus strain selected for each qPCR design, the position of the target sequence on the selected viral strain and the amplicon size.

| **Institution** | **qPCR design** | **Coronavirus selected** | **Position in coronavirus selected** | **Amplicon size** |
| --- | --- | --- | --- | --- |
| Institut Pasteur<br>(Institut Pasteur, 2020) | IP2 | Pangolin CoV - MT121216.1 | 12549 – 12656 bp | 108 bp |
|  | IP4 | Pangolin CoV - MT121216.1 | 13939 – 14045 bp | 107 bp |
| Charité<br>(Corman et al., 2020) | RdRp-P2 | Pangolin CoV - MT121216.1 | 15290 – 15389 bp | 100 bp |
|  | E | Pangolin CoV - MT121216.1 | 26103 – 26215 bp | 113 bp |
| CDC Atlanta<br>(CDC Atlanta, 2020) | N1 | Pangolin CoV - MT121216.1 | 28073 – 28144 bp | 72 bp |
|  | N2 | Pangolin CoV - MT072864.1 | 29102 – 29166 bp | 65 bp |

These results agree with those recently published by Liu et al. (2020) and MartÍnez-Murcia et al. (2020), where Bat CoV RaTG13 and Pangolin CoV sequences showed the highest similarities with SARS-CoV-2.

As illustrated in the alignment of the qPCR designs (primers and probes) and their closest Pangolin CoV sequences (Fig. 1), both IP2 and IP4 showed 4 mismatches, 2 located in probe and 2 in reverse primer; N1 showed 2 mismatches, 1 in each primer; N2 showed 4 mismatches, 1 in forward primer and 3 in probe; RdRp-P2 showed 3 mismatches, 1 in each primer and 1 in probe; E showed full matching; and finally GPS™ *CoVID-19 dtec-RT-qPCR Test* showed a total of 21 mismatches, with the highest number of them located in reverse primer. According to these, we considered the possibility that IP2, IP4, N1, E and RdRp-P2 may amplify samples which would contain strains of selected Pangolin CoV, or other non-described viral strains with similar sequences. Therefore, to ascertain this prediction based on the *in silico* analysis with an *in vitro* test, was the purpose of this study. As the E qPCR design showed full matching with the selected coronavirus sequence, its corresponding *in vitro* test was excluded from this analysis. Nevertheless, the qPCR designed for gene E was conceived to be inclusive for virus of the subgenus *Sarbecovirus* (Corman et al., 2020).

**Figure 1.**
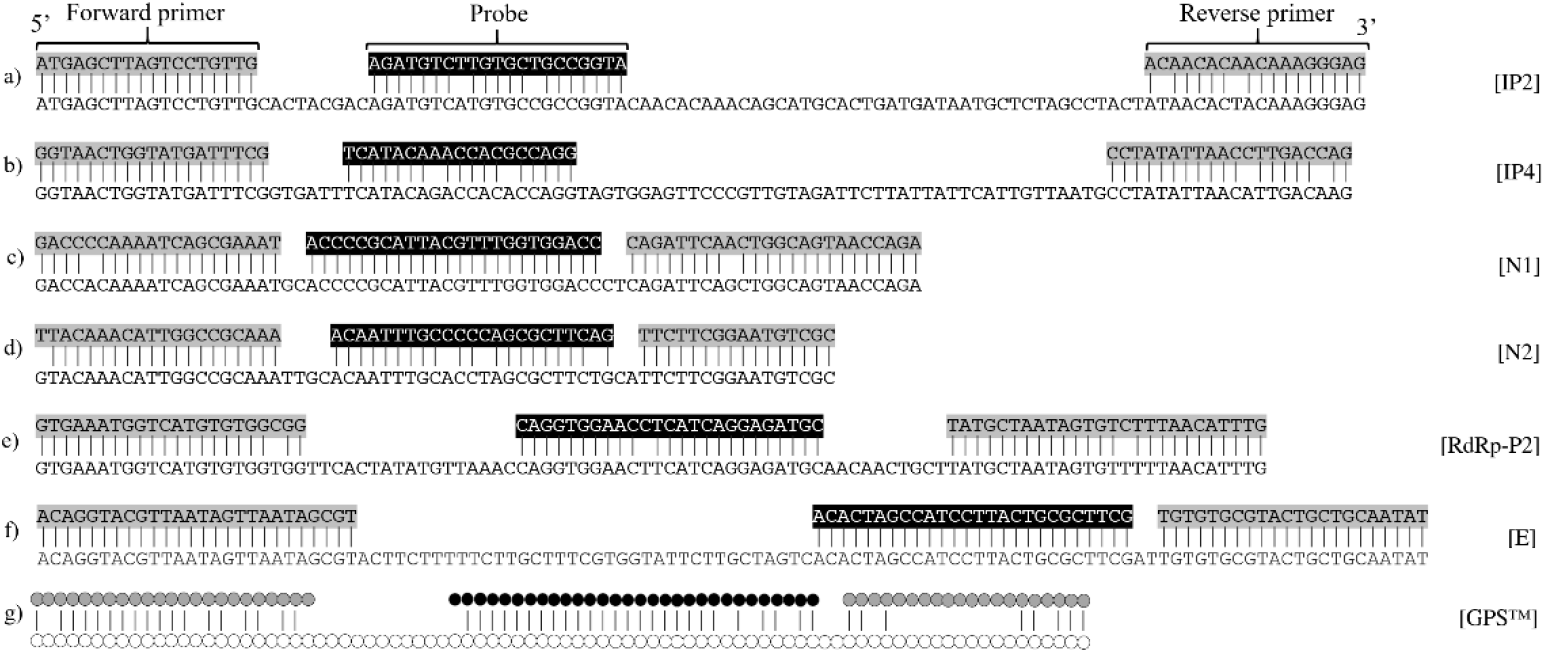
Sequence alignment between primers and probes of: a) IP2 with Pangolin CoV - MT121216.1; b) IP4 with Pangolin CoV - MT121216.1; c) N1 with Pangolin CoV -MT121216.1; d) N2 with Pangolin CoV - MT072864.1; e) RdRp-P2 with Pangolin CoV - MT121216.1; f) E with pangolin CoV - MT121216.1; g) GPS™ *CoVID-19 dtec-RT-qPCR Test* with Pangolin CoV - EPI_ISL_410539.

Experimental work to test the *in vitro* exclusivity of all these qPCR designs was carried out by testing each template (10 to 10^6^ copies) of the Pangolin CoV selected sequences, and results are shown in Figure 2. Positive controls for all qPCR tests were also performed by using synthetic templates (10 to 10^6^ copies) with sequences showing full-matching to the corresponding qPCR design, and they all gave exponential amplification (Fig. 2). The results indicated that IP2, N1 and RdRp-P2 qPCR designs were able to amplify the nucleotide sequence of the selected Pangolin CoV strains, which validates the hypothesis of possible false positives when testing exclusively for SARS-CoV-2. In the cases of N1 and RdRp-P2 (with 2 and 3 mismatches, respectively), Ct values obtained were very close to this of the corresponding positive controls, which suggests a hybridization relatively stable of oligonucleotide primers and probes with target sequences. However, the IP2 qPCR design, showing 4 mismatches, yielded Ct values considerably delayed from these of the positive control; thus, it seems this amplification, although it occurred, was much weaker. IP4 and N2, both showing 4 mismatches, showed no qPCR amplification at all; so, in these two cases, mismatching seems to be more efficient discriminating closely related sequences. The experiments were repeated 3 times and results were reproducible in all cases. End-point PCR was assessed with primers of IP4 and N2 (excluding the corresponding probes) and very robust bands of the expected fragment-size were obtained in the agarose gels (not shown). This finding suggests that amplification may occur in the qPCR, but the discriminative character may reside in the probe (2 and 3 mismatches, respectively). As expected, the GPS™ kit showed no amplification when tested against the most similar described sequence (21 mismatches). It should be noted that the GPS™ kit design undergoes continuous revision to ensure its full specificity with respect to all the variants that are emerging as a consequence of the high mutation rate of the SARS-CoV-2.

**Figure 2.**
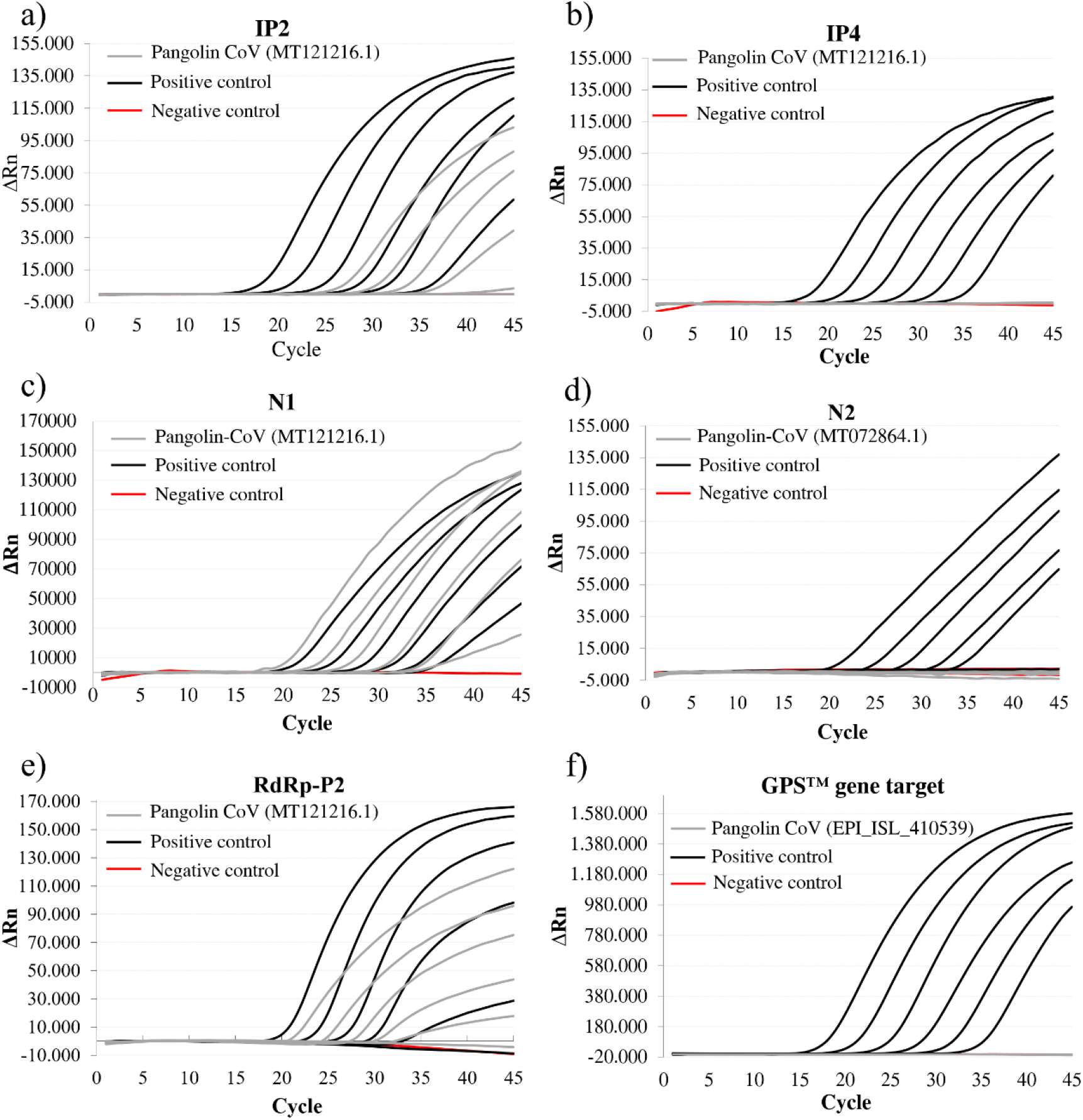
Amplification plots of synthetic DNA template dilutions from 10 to 10^6^ copies of positive controls and Pangolin CoV nucleotide sequences, and negative controls, using the a) IP2; b) IP4; c) N1; d) N2; e) RdRp-P2; f) GPS™ *CoVID-19 dtec-RT-qPCR Test* qPCR designs.

These results demonstrate that some qPCR designs may be able to detect SARS-CoV-2-related coronavirus, such these from Pangolin CoV strains, and maybe similar *Sarbecovirus* non-described yet. This finding may do not have relevance on human diagnostics for clinical specimens, as currently, animal-specific coronavirus should not be frequently found in human samples. Additionally, the most common human coronavirus strains which cause breathing illnesses, like OC43, HKU1, NL63 and 229E, show a low homology with SARS-CoV-2 (Zhou et al., 2020). However, the exclusivity of the qPCR design is particularly relevant for SARS-CoV-2 monitoring in environmental samples, for instance in the epidemiological surveillance of wastewaters. Several laboratories have developed protocols for analysis of water samples by using some of these published qPCR designs to detect SARS-CoV-2 (Chavarria-Miró et al., 2020; Fongaro et al., 2020; Randazzo et al., 2020b), evaluated in the present study. Some of their results, although with a very low signal, suggest the presence of SARS-CoV-2 in wastewater samples harvested in countries remotely distant from China, much time before (12^th^ March 2019 and 27^th^ November 2019) the first outbreak was formally detected (31^th^ December 2019). Above finding was supported by very low qPCR signals (Ct values ranging 39-40, Chavarria-Miró et al., 2020). Having ruled-out a possible cross contamination of PCR reactions, some of these weak-positives may be explained by the consequences recruited in this work. They may come from other highly similar coronavirus sequences, rather than SARS-CoV-2.

Considering the current knowledge about the huge extent of microbial diversity still to be described (number of cultured/described species have been estimated to represent ca. 4 – 5% from this existing in nature), it is crucial to take into account that environmental samples may contain a pool of many different but closely related viruses, consequently a considerable number of different sequences were unknown at the time of qPCR design. The use of highly exclusive primers and probes to detect only the SARS-CoV-2 causing CoVID-19 may help on the purpose but, definitely, studies to discriminate this from the real diversity should be complemented by using massive sequencing techniques, like next-generation sequencing (NGS).

## Data Availability

All data referred to in the manuscript is available.

## Ethics Statement

The authors confirm that the ethical policies of the journal, as noted on the journal’s author guidelines page, have been adhered to. No ethical approval was required.

